# *Screen Your Way* Study Protocol: Embedding community driven models to increase cervical screening via HPV self-collection to improve cervical cancer outcomes for Aboriginal and Torres Strait Islander people

**DOI:** 10.1101/2025.10.21.25338491

**Authors:** Lisa J Whop, Louise E Mitchell, Tamara L Butler, Deborah Wong, Kate Wilkinson, Joan Cunningham, Sonya Egert, Kristine Falzon, Gail Garvey, Rebecca Guy, Beverley Lawton, Hamish McManus, Claire Nightingale, Marion Saville, Megan Smith, Claudette “Sissy” Tyson, Mark Wenitong, Claire Zammit, Karen Canfell, Natalie Taylor, Julia Brotherton

**Author notes:** Corresponding author:, Australian New Zealand Clinical Trials Registry Number: ACTRN12625001134415.

## Abstract

In July 2022, Human Papillomavirus (HPV) self-collection became available as a choice to all participants in Australia’s National Cervical Screening Program (NCSP). This policy change aims to facilitate equitable access to cervical screening; however, further evidence is needed to support its implementation and reach under-screened women and people with a cervix. This implementation study aims to embed HPV self-collection into Aboriginal and Torres Strait Islander Community Controlled Health Organisations (ACCHO) and/or primary care organisations whose context is similar to that of an ACCHO. This will be achieved by co-designing, implementing, and evaluating models of care tailored to local needs. The aim is to increase cervical screening participation, particularly among under- and never-screened, Aboriginal and Torres Strait Islander women and people with a cervix. Ultimately the aim is to achieve equity in cervical cancer elimination.

*Screen Your Way* will use a before-and-after study design to evaluate the effectiveness, acceptability and sustainability of implemented strategies on cervical screening participation among Aboriginal and Torres Strait Islander women and people with a cervix. The study will be guided by an Indigenist research approach and will employ mixed methods.

Ethical approval has been obtained from the Australian Institute of Aboriginal and Torres Strait Islander Studies Research Ethics Committee (REC-0092), Aboriginal Health and Medical Research Council of New South Wales Ethics Committee (2078/23), the Australian National University Human Research Ethics Committee (H/2023/1103), and the Northern Territory Department of Health and Menzies School of Health Research (HREC2023-4557). Additional ethical approvals will be obtained as required by individual ACCHOs. Findings will be disseminated via workshops, reports, evidence briefs and resource creation to assist with the evidence-based scale up of self-collection in the ACCHO setting. Further dissemination will occur via conferences and peer-reviewed publications in partnership with the Screen Your Way Aboriginal and Torres Strait Islander Caucus.

## Introduction

Cervical cancer is preventable and, if detected early and managed effectively, can be successfully treated. (1, 2) This understanding prompted Australia to adopt a nationally coordinated approach to prevention of cervical cancer through the National Human Papillomavirus (HPV) Vaccination Program, implemented in 2007, and the National Cervical Screening Program (NCSP), implemented in 1991. Since the inception of these programs, Australia has seen a 50% reduction in cervical cancer incidence, (3) and is ‘on track’ to become the first country to eliminate cervical cancer. (2, 4) However, inequities persist and without targeted action, elimination is unlikely to include Aboriginal or Torres Strait Islander women and people with a cervix. (5) Cervical cancer burden among Aboriginal and Torres Strait Islander women and people with a cervix remains unacceptably high. After adjusting for age, cervical cancer incidence was 2.3 times higher in 2016-2020 and mortality was 3.6 times higher in 2018-2022 among Aboriginal and Torres Strait Islander women and people with a cervix when compared to non-Indigenous people of screening age. (6)

In December 2017, the NCSP transitioned from cytology based (Papanicolaou or “Pap” test) to HPV DNA based primary screening, facilitating the introduction of a self-collection screening pathway. Self-collection allows for women and people with a cervix to take their own vaginal sample using a flocked swab to test for the presence of HPV. Initially, evidence suggested that self-collection was associated with a small loss in sensitivity compared to clinician-collected samples. (7) Initially, self-collection was offered within the NCSP under a restricted model. It was available to under- or never-screened women and individuals with a cervix aged 30 years or older who had declined a clinician-collected test. These restrictions led to multiple implementation barriers, including poor uptake by clinicians and low awareness among eligible participants. (8) In the first five years of its availability, only 6,000 under-screened women and people with a cervix participated in self-collection, representing less than 1% of the estimated 1 million eligible unscreened individuals. (9) The number of Aboriginal and Torres Strait Islander participants who used self-collection is unknown. At that time, few Aboriginal and Torres Strait Islander women and people with a cervix were aware of self-collection and many identified the need for support to complete it confidently and safely. (10) In July 2022, based on strong evidence of the equivalent sensitivity between self-collection and practitioner-collection, Australia updated its policy to make self-collection available to all cervical screening participants. (11, 12) This change aimed to remove some of the barriers impacting cervical screening participation and reduce inequities in cervical cancer incidence and mortality. (13)

To reach the elimination threshold, cervical cancer incidence must be reduced by 67% based on 2015-2019 incidence rates for Aboriginal and Torres Strait Islander women and people with a cervix. (5, 14) This requires a significant increase in screening among under- and never-screened individuals. Nationally, cervical screening coverage estimates were 73.1% of the eligible population under the renewed program from 2019-2023. However, there are remoteness and socioeconomic gradients, with lower rates of screening among those living remotely or experiencing socioeconomic disadvantage. (6)

Additionally, unacceptable disparities persist in cervical screening rates and outcomes for Aboriginal and Torres Strait Islander women and people with a cervix. While data by Indigenous status is not routinely available from the NCSP, localised studies consistently show that participation in cervical screening is substantially lower for Aboriginal and Torres Strait Islander women compared to other women, averaging 20 percentage points lower. (15, 16) Even within Aboriginal Community Controlled Health Organisations (ACCHO), which provide a supportive and culturally safe healthcare setting, only 42% of screen-eligible Aboriginal and Torres Strait Islander regular clients had a cervical screening test recorded in the local patient management system within the previous five years as of June 2023. (17)

To achieve cervical cancer elimination for all women and people with a cervix, including priority groups such as Aboriginal and Torres Strait Islander peoples, Australia must urgently address systemic failures. Significant access barriers impact the ability of Aboriginal and Torres Strait Islander women and people with a cervix to access cervical screening. Invasive clinical examinations have long been a barrier to participating in cervical screening. (10, 18) Other barriers to self-collection include difficulties in identifying eligible participants, interpreting guidelines, and implementation issues limiting primary care providers ability to offer self-collection. (8, 19) These barriers, combined with distrust of colonial institutions, legacies of intergenerational trauma and racism experienced within mainstream healthcare settings, necessitate concerted efforts to improve the experience of cervical screening for Aboriginal and Torres Strait Islander women and people with a cervix. (5, 10) Strategies that prioritise Indigenous knowledge and strengths, without reinforcing misconceptions that fuel racism and inequity within the Australian health system, are essential.

The availability of universal self-collection provides an opportunity to address inequities in cervical screening among Aboriginal and Torres Strait Islander women and people with a cervix. Research suggests that Indigenous women in westernised, settler-colonial countries find HPV self-collection highly acceptable, as it increases privacy, comfort, convenience, power and control, which are integral to participation. (10, 20–22) The major concerns regarding self-collection related to correctly administering the test and its accuracy. However, Australian research shows broad satisfaction with the instructions (22, 23) and low rates of unsatisfactory test returns. (24) While NCSP data is not yet available for Aboriginal and Torres Strait Islander women and people with a cervix, research indicates that self-collection is highly acceptable to Indigenous peoples globally, significantly increasing their cervical screening participation in both trial and organised screening program settings. (23–27)

The next step is to sustainably embed self-collection within the ACCHO context. In 2018-2019, 48% of Aboriginal and Torres Strait Islander people indicated they would prefer to access primary care via an ACCHO (390,600 people) and 34% accessed one as their usual source of primary care (277,100 people). (28) Supporting Aboriginal and Torres Strait Islander women and people with a cervix to access self-collection in the culturally safe environment of an ACCHO should translate into an actual increase in cervical screening participation and be a major contributor to achieving equity in cervical cancer elimination.

### Rationale and aims

Using Rigney’s Indigenist Research approach (29) and guided by the Lowitja Institute Evaluation Framework, (30) this study aims to increase cervical screening participation among under- and never-screened Aboriginal and Torres Strait Islander women and people with a cervix in partnership with ACCHOs by implementing HPV self-collection into practice.

The objectives of *Screen Your Way* are to:

1. Co-design, implement and evaluate models to support HPV self-collection and other strategies that increase cervical screening within ACCHOs
2. Assess the impact of HPV self-collection implementation models on cervical screening participation
3. Explore providers’ and clients’ views and experiences of the project to provide context for the cervical screening participation data and inform the evaluation of the implementation strategies
4. Disseminate findings to assist with the evidence based national scale up of HPV self-collection.

## Materials and Methods

*Screen Your Way* will use a before-and-after study design to evaluate the effectiveness of strategies on cervical screening participation among under-screened Aboriginal and Torres Strait Islander women and people with a cervix during the study period. This protocol has been developed following the Standard Protocol Items: Recommendations for Interventional Trials (SPIRIT) checklist (see Fig 1 and Supplementary Information). (31) The study was retrospectively registered on 16/10/2025 in the Australian New Zealand Clinical Trials Registry (ANZCTR; ACTRN12625001134415, https://anzctr.org.au/ACTRN12625001134415.aspx. The study was initially assessed as not requiring prospective trial registration. Registration was subsequently completed. The authors confirm that all ongoing and related trials for this intervention are registered.

**Fig 1.**
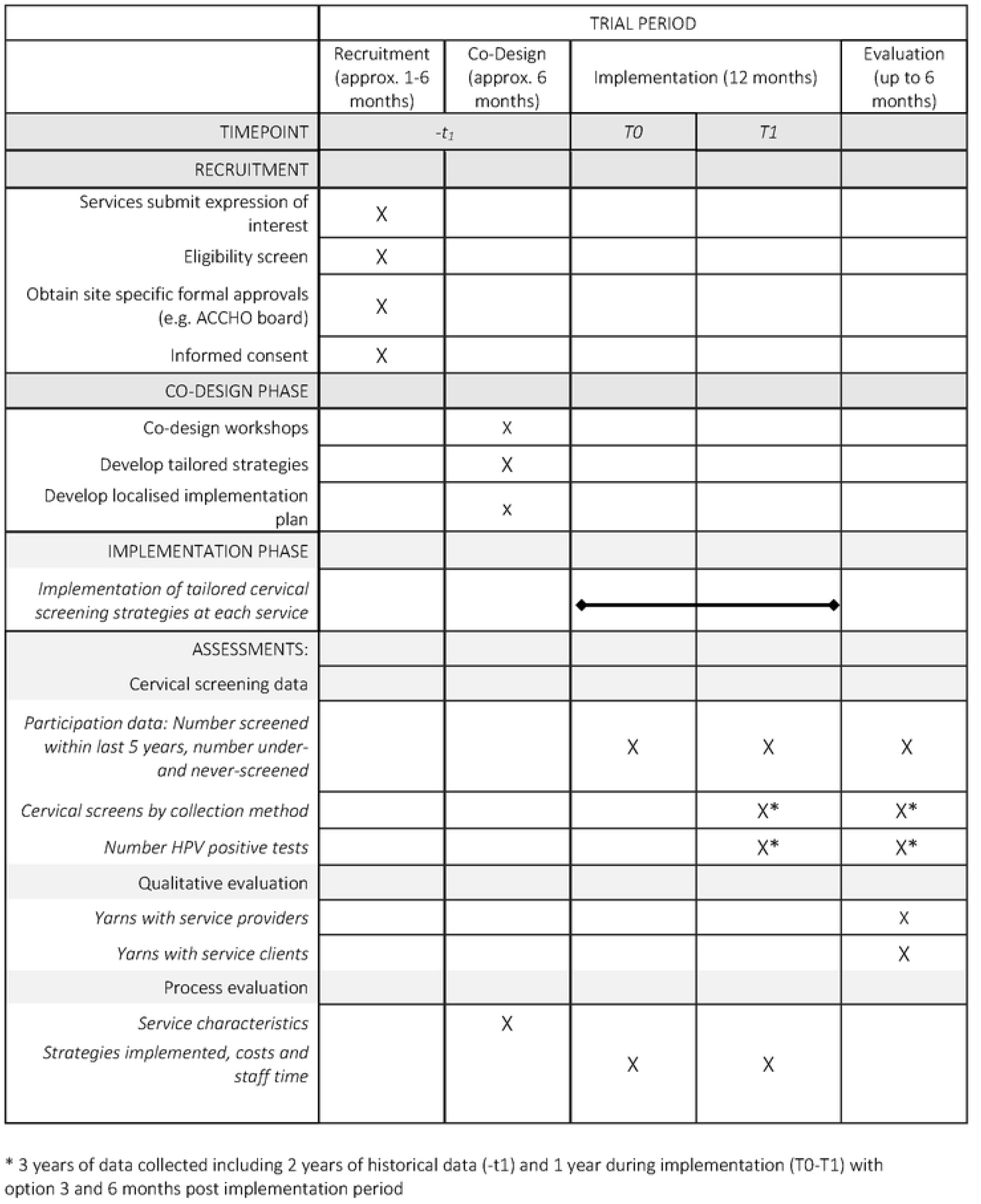
SPIRIT schedule of recruitment, co-design, implementation and evaluation phase for Screen your Way.

### Approach and guiding principles

*Screen Your Way* is designed to deliver equitable, sustainable and scalable reductions in cervical cancer incidence through cervical screening by fostering strong relationships between communities and organisations; their combined local expertise and priorities will shape the research design and implementation. This project takes a systems approach, recognising that Aboriginal and Torres Strait Islander women and people with a cervix have various needs, within and outside of the health system. The theory of change is illustrated in Fig 2.

**Fig 2.**
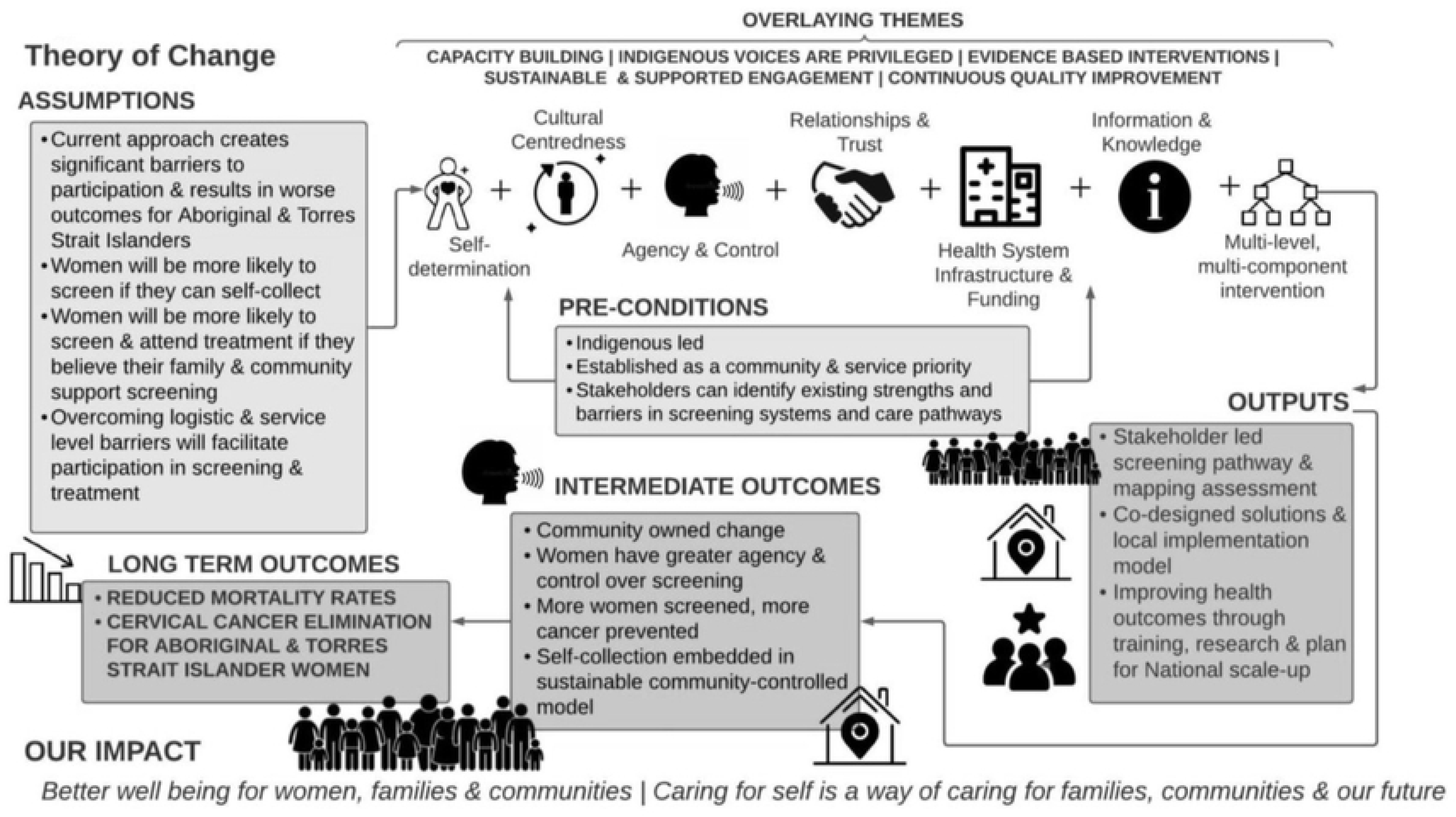
Screen Your Way: Theory of Change Model.

A multidisciplinary mixed-methods research approach will be used that incorporates Indigenous research methodologies, foregrounds Indigenous intellectual sovereignty and leadership, and is implementation focused. The program of research is grounded within Rigney’s Indigenist Research Approach (29) which will then be designed, implemented and evaluated in accordance with the principles outlined within the Lowitja Evaluation Framework (30), with a key focus on identifying elements important to implementation success in Indigenous communities (e.g. the criticality of Indigenous self-determination, concept of culture centredness, community control and, in primary care, quality improvement processes). (32)

### Governance

The Governance Framework illustrated in Fig 3 will ensure cultural and clinical oversight of the research and maximise real-time knowledge sharing. Cultural governance will be monitored by Aboriginal and Torres Strait Islander Reference Group, Thiitu Tharrmay, at the Australian National University. Thiitu Tharrmay in Ngiyampaa language translates as ‘to share/exchange knowledge’. An Aboriginal and Torres Strait Islander Project Caucus comprised of Aboriginal and Torres Strait Islander Chief and Associate Investigators will provide over-arching project governance and ensure that the project is culturally safe and meets the needs and priorities of Aboriginal and Torres Strait Islander people and communities. An Investigator team has been established in line with the Medical Journal of Australia guidelines (33) and the principles of the Indigenist research approach. (29, 34)

**Fig 3.**
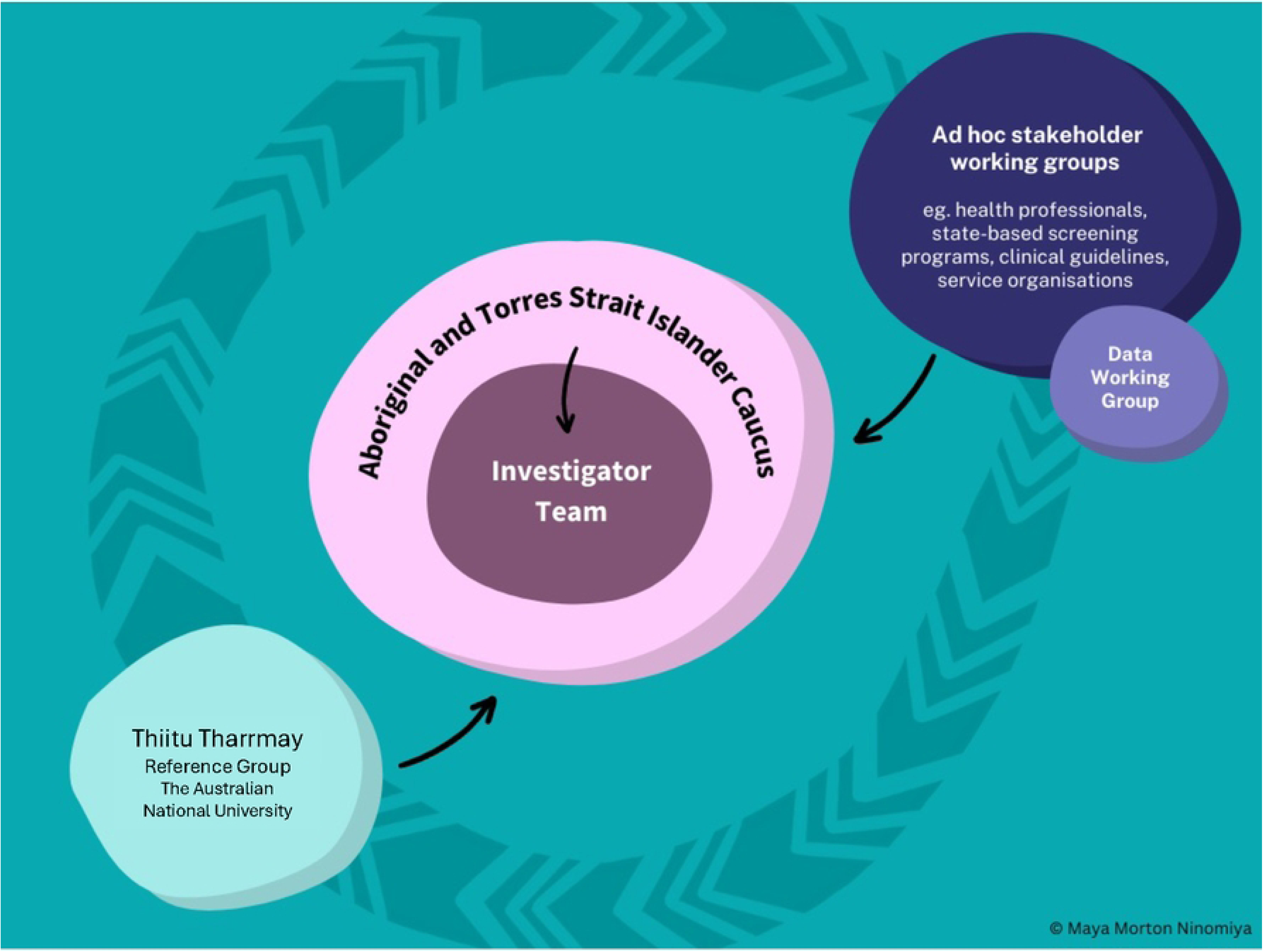
Screen Your Way Governance Framework.

### Setting

*Screen Your Way* will take place in ACCHOs and/or primary care organisations whose context is similar to that of an ACCHO in the Australian States and Territories of Queensland (Qld), New South Wales (NSW) and the Northern Territory (NT). At the last Census date more than 164,000, or over 70%, of Aboriginal and Torres Strait Islander women of screening age were recorded to live across these three jurisdictions. (35) Sampling in this setting will provide geographical and service diversity in the study to inform national scale up plans. Service recruitment commenced on the 1st October 2023, when ACCHOs were invited to submit an expression of interest to participate in the study. Services were recruited between 5^th^ February 2024 and 8^th^ August 2025. Following recruitment, each site undertook a co-design phase tailored to local needs. The 12-month implementation period began at sites in between the 1^st^ September 2024 and is expected to continue until 8^th^ September 2026, depending on each site’s start date. Data collection is anticipated to be completed by 30^th^ September 2026, with study results expected in 2027.

### Study Design

After recruitment of services to the study, strategies will be co-designed with each service to meet local needs and then implemented for 12 months to allow for a maximum number of screening events. An overview of the study design can be found in Fig 4.

**Fig 4.**
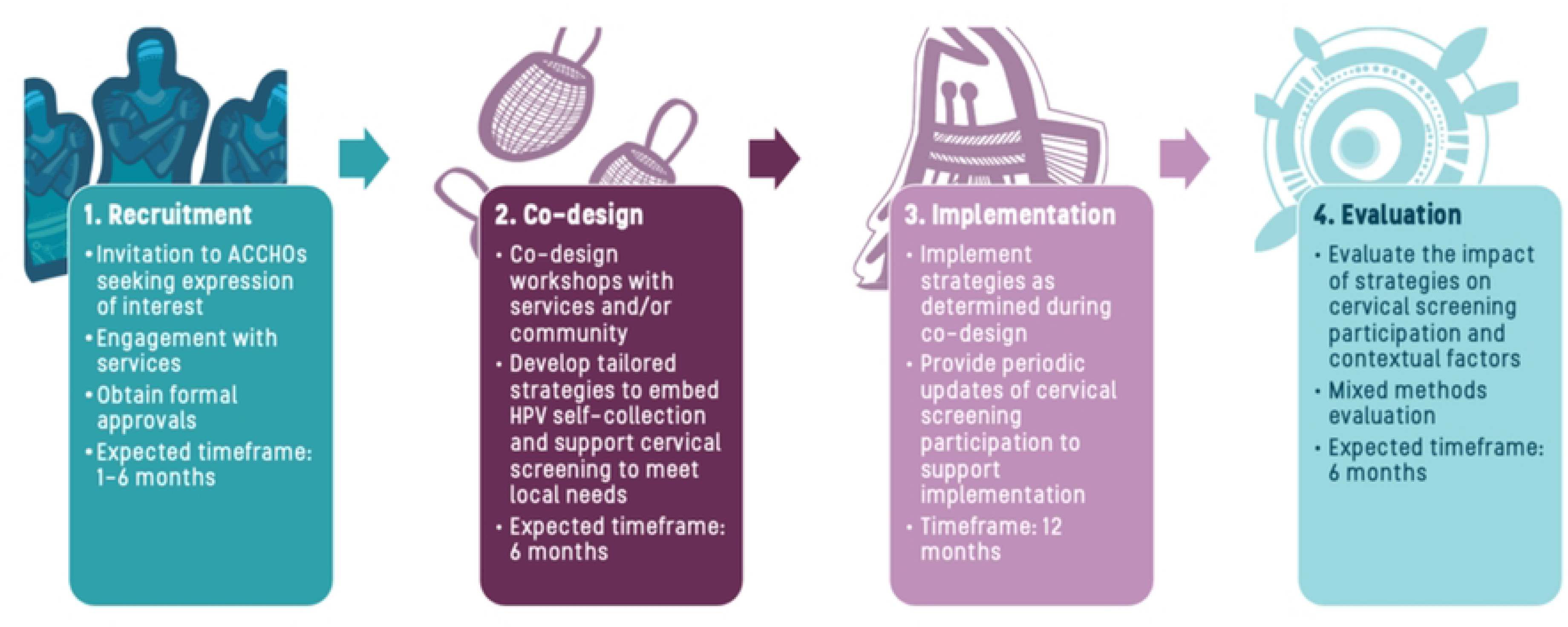
Steps for working with Aboriginal Community Controlled Health Organisations.

### Step 1: Recruitment of services to participate in *Screen Your Way*

A minimum of six services (two per jurisdiction) will be recruited, using a process similar to that used successfully with ACCHOs in the literature. (36, 37) Support to approach services about participation in the research has been obtained from relevant community-controlled National and State-based governing bodies, including National Aboriginal Community Controlled Health Organisation (NACCHO), Queensland Aboriginal and Islander Health Council (QAIHC), Aboriginal Health and Medical Research Council (AHMRC) of NSW and Aboriginal Medical Services Alliance Northern Territory (AMSANT). Services will be invited to submit an expression of interest. The communication avenue(s) used will through existing networks and guided by QAIHC, AMSANT and AHMRC but will likely include contact through their member email lists and newsletters (usually sent to service Chief Executive Officers, Medical Directors and/or Boards).

Services will be purposefully sampled in line with the following eligibility criteria:

1. Are an ACCHO, or a primary healthcare organisation with a majority of clients being Aboriginal and Torres Strait Islander people whose context is similar to that of an ACCHO (referred to as ACCHOs hereinafter)
2. Are located in Qld, NSW or the NT
3. Have 120 or more Aboriginal and Torres Strait Islander women or people with a cervix aged 25-74 years who are active clients (defined as three or more clinic visits in past two years)
4. Have at least one cervical screening certified health professional
5. Consent to provide screening data related to cervical screening.

Services will be excluded if they are unable to meet the inclusion criteria or if they are involved in other trials relating to cervical screening given the potential to contaminate results.

Written informed consent will be obtained from the ACCHO Chief Executive Officer, Medical Director, and/or Board. Consent may be withdrawn from the study at any time before dissemination of the findings. Individual patient consent for this study will not be sought from clients attending the health service for the provision of deidentified cervical screening data or for the implementation of strategies as these activities would be part of routine cervical screening occurring at the service. Where assistance is requested to support data cleaning or audit, additional ethics approval will be sought for research personnel to access client level data for this purpose. Individual informed consent will be obtained for Yarns at the time of participation.

### Step 2: Co-design of strategies for implementing self-collection at each site

We will work with nominated staff at participating services to facilitate co-design workshops, the design of locally tailored strategies and to engage with community members as determined locally in line with keys principles of co-design with Aboriginal and Torres Strait Islander peoples including leadership, a culturally grounded approach, respect, benefit to community, inclusive partnerships and transparency and evaluation and reciprocity. (38) (39) Co-design will be tailored to the local context with study personnel offering two visits.

The aim of the strategies is to increase the participation of under-and never screened women and people with a cervix in cervical screening using HPV self-collection. Strategies developed during the co-design process will be tailored to meet the local service needs and the communities they serve. Strategies span community engagement, participant level, workforce level, and system and/or health service level strategies.

By co-designing within an Indigenist Research Approach, we ensure that Aboriginal and Torres Strait Islander people are the ‘*architects of health advancement rather than accessories to failed health policy frameworks*’ (page 198). (40) Participating services will work closely with the research team to create personalised evidence-based implementation models suited to their service and the communities they serve. This step is anticipated to take approximately six months.

### Step 3: Implementation of cervical screening strategies

Participating services will be provided a customised report with findings from co-design workshops. This will be used to clearly define the strategies chosen and outline the roles and responsibilities of the research team and the service in implementing the strategies. Support will be provided to update cervical screening data (as requested) along with direct research costs to further promote availability of self-collection and cervical screening during the pilot period, using the most appropriate methods for their community. The research team will undertake resource development, facilitate training of the workforce and other means of strengthening existing capabilities in cervical screening as determined during co-design. Services will implement the co-designed strategies for 12 months to improve delivery of cervical screening in their communities and increase service level participation. Strategies will be implemented with support from research staff tailored to local service requests.

### Step 4: Evaluation

The evaluation of this research will be conducted within an overarching conceptual framework that combines key constructs from the RE-AIM Framework (41) with the Health Equity Implementation Framework. (42) The evaluation variables are outlined in Table 1. During the co-design process services may identify additional variables or research questions of interest to their unique context and these will be added to the evaluation framework.

**Table 1.** Evaluation variables.

| Approach | Collection methods | Variables |
| --- | --- | --- |
| Quantitative | De-identified data extracted from service Record Management Software and/or Clinical Audit Tools at baseline (#T0), periodic intervals as determined by each service (approximately 3 monthly), and 12 months for pre-post analysis (#T1). Services can provide data at 3 and 6 months post implementation. | De-identified cervical screening data for screen eligible Aboriginal and Torres Strait Islander clients at each service (number and proportion):<br>Cervical screening participation #T0, #T1<br>Under-or never-screened #T0, #T1<br>Cervical screens by collection method (i.e. self- or clinician-collection) #T1<br>HPV positive tests #T1 |
| Qualitative | Qualitative Yarns and Yarning circles with ACCHO staff and eligible clients (women and people with a cervix) | Yarns will cover topics such as: experience of participating in or implementing strategies; perceptions of effectiveness; reach and sustainability of strategies; challenges and strengths throughout implementation period. |
| Process | Baseline survey completed by staff<br>Worksheet to complete process data<br>Yarns with service providers | Service characteristics (e.g. workforce)<br>Number and type of strategies implemented<br>Costs and staff time to implement strategies<br>Barriers to adoption and sustainability |

### Data Collection and analysis

#### Cervical Screening Data

The primary outcome will be the number and proportion of Aboriginal and Torres Strait Islander women and people with a cervix who are overdue for cervical screening (under-screened) or have no recorded history of a cervical screen (never-screened) at the completion of the study, compared to the before period. Secondary outcomes are cervical screening participation, the number and proportion of cervical screens conducted by collection type (self-collect versus clinician collect) and HPV positive tests in the 2-years before and during the study period. Quantitative outcomes data will be obtained using one or a combination of the following pathways to be determined by the services’ preferences: from local practice management software, practice audit tools, from the National Cervical Screening Register (NCSR) and/or manual audit of relevant patient records. De-identified data will be provided to nominated members of the research team via secure methods.

Descriptive analysis will be used to analyse cervical screening data. Primary and secondary outcomes will be calculated comparing data from the before (*T0*) and after (*T1*) period. Retention of effects will be measured for services who provide data at 3- and 6-month intervals in the post-implementation period. For the primary outcome, the number and proportion of all screen eligible clients at the services who are under-screened or never-screened at *T1* compared to *T0* (number of under- and never-screened screen eligible clients/number of screen eligible women). Secondary outcomes are cervical screening participation rate (number with a recorded cervical screen in the previous 5-years/number of screen eligible clients), and the number and rate of cervical screens conducted using self-collection (number of tests conducted using self-collection/number of screening tests), and the HPV positivity (number of HPV positive tests/number of screening tests).

#### Yarns

Semi-structured Yarns and Yarning circles will be used to explore providers’ and clients’ views and experiences of the project, evaluate implementation strategies and to provide contextual information to support cervical screening participation data. Data collection will include individual and group Yarns with approximately 20-30 service staff involved in cervical screening strategies (including doctors, nurses, and Aboriginal Health Practitioners) and separately with 20-40 women and people with a cervix who are clients of the service and who were offered cervical screening. All participants (both screening participants and providers) will provide individual informed consent prior to the Yarns or Yarning circles. Yarns may take place any time during the implementation (after) period of the study but will be concentrated towards the end of, or after, the 12 months allocated to the implementation of strategies. Yarns may take place in-person, online via virtual meeting software or over the phone and will be audio-recorded and transcribed verbatim.

Yarning is a culturally appropriate Indigenous research methodology grounded in Aboriginal and Torres Strait Islander peoples’ traditional ways of communicating, building relationships and sharing knowledge. (43, 44) It involves distinct but interconnected types of Yarning including social Yarning to establish relationality and trust, research Yarning to explore the research topic, and therapeutic Yarning to create space for healing conversations if needed. (43) Where possible, trained female Aboriginal and/or Torres Strait Islander researchers will Yarn with participants. Non-Indigenous researchers may also conduct Yarns with appropriate training and mentoring from Aboriginal and Torres Strait Islander people to ensure a culturally safe approach.

Data (transcripts and field notes) will be managed with NVivo software (QSR International, V.12). Framework analysis (also known as “codebook thematic analysis” (45)) will be used to systematically identify themes in the data, using both inductive and deductive coding. (46) In line with the Indigenist research approach, Aboriginal and Torres Strait Islander people will lead analysis, with support from non-Indigenous researchers. This approach centres Aboriginal and Torres Strait Islander people’s worldviews and lived experiences in the interpretation and analysis of the data. The Lowitja Evaluation Framework will guide analysis, ensuring social, structural and environmental factors influencing the uptake of self-collection are considered alongside individual level factors and grounded within an Indigenist research approach.

#### Process data

Process data will be used to examine the fidelity of strategy implementation and explore factors that may have influenced their effectiveness in increasing cervical screening participation. Process data will include service level information (including the service size, number of eligible clients, the number of cervical screening providers and provider type, remoteness; as well as information on the range of activities implemented in the models e.g. resources/materials used, provider training activities, community events undertaken, location of self-collect, recalls set, reports provided. Any unrelated activities that may have influenced health care attendance (e.g., major cultural or other health promotion activities) will be captured. A worksheet based on the Framework for Reporting Adaptations and Modifications to Evidence-based Implementation Strategies (FRAME-IS) (47) will be used to collect process data. This tool will allow the research team and the ACCHOs to track when, why and how strategies have been implemented and any associated costs or time.

### Ethics

This project respects and acts in accordance with the ethical guidelines set out by the Australian Institute of Aboriginal and Torres Strait Islander Studies (AIATSIS) to ensure respect, integrity, and collaboration in all our work. (48) Screen Your Way is Aboriginal and Torres Strait Islander designed, led and governed. Approval has been obtained from the AIATSIS Research Ethics Committee (REC-0092), the Aboriginal Health and Medical Research Council of New South Wales Ethics Committee (2078/23), the Australian National University Human Research Ethics Committee (H/2023/1103), and the HREC of the Northern Territory Department of Health and Menzies School of Health Research (HREC 2023-4557). Additional approvals will be obtained to meet local ACCHO requirements.

## Discussion

### Dissemination Plan

Our co-design process will develop materials, procedures and processes which can be used broadly, beyond the research. Our partnership and governance structure will ensure efficient knowledge transfer mechanisms, facilitated through respectful collaboration, including leaders in the Aboriginal community-controlled sector, cervical screening and academia. Findings will be disseminated to support the evidence-informed national scale up of self-collection. This will include publication in peer review journals and presentations at conferences. Publications will adhere to CONSolIDated critERtia for strengthening the reporting of health research involving Indigenous Peoples (CONSIDER) statement. (49) Findings will also be shared in a variety of other formats previously shown to be successful in translating implementation research findings into policy and practice including translational workshops, evidence briefs and presentations, reports and the creation of resources.

### Implications

This research directly aligns to recommendations in Australia’s National Strategy for the Elimination of Cervical Cancer (50) and the World Health Organization’s Global Strategy to Accelerate the Elimination of Cervical Cancer (2) where the immediate priority is: to save women’s lives in the short term by scaling up screening participation and receipt of treatment globally by 2030. Overcoming universal self-collection implementation issues will be necessary to increase participation and achieve equity in cervical cancer elimination for Aboriginal and Torres Strait Islander people.

Co-designed strategies have the potential to transform practice and outcomes, and study findings will be readily transferrable to ACCHOs across Australia. They are also likely to be adaptable to Indigenous women and people with a cervix in other countries. By encouraging community-driven innovations relating to self-collection, we aim to embed these practices in ACCHO service delivery long after the study is completed and provide a way to take the findings and implement nationally. This will both honour community voices and maximize cervical screening participation through a mechanism that we know works. This is critical to ensure that Aboriginal and Torres Strait Islander women and people with a cervix are not left behind as Australia approaches elimination of cervical cancer.

## Data Availability

No datasets were generated or analysed during the current study (study protocol). In line with Indigenous Governance and ethical requirements of health service data this data will not be made openly available upon study completion.

## Authors contributions

LJW conceived the initial study design. All authors contributed to the conception, design, and development of this protocol. Each author participated in drafting the manuscript, revising it critically for important intellectual content, and approving the final version to be published. All authors agree to be accountable for all aspects of the work.

## Acknowledgements

For the artwork, we acknowledge and thank Simone Arnol and Bernard Lee Singleton, Yalma. Ownership of Aboriginal and Torres Strait Islander knowledges and cultural heritage will be retained by the informants. We also thank the contribution of Thiitu Tharrmay members.

## Supporting Information

S1. SPIRIT Checklist

## Financial Disclosure Statement

This research has been funded through an Australian National Health and Medical Research Council (NHMRC) Targeted Call for Research competitive funding grant (GNT201490). LJW is supported by a NHMRC EL2 Investigator grant (2009208); TB is supported by a NHMRC Investigator Grant (2008097) EL1; JC is supported by NHMRC Research Fellowship (1058244); GG is supported by NHMRC Investigator grant L2 (1176651). The funding organisation will have no role in the collection, management, analysis, and interpretation of data; writing of the protocol or subsequent manuscripts; and the decision to submit the protocol for publication. The views expressed in this publication are those of the authors and do not necessarily reflect the views of the funders.

## Competing interests

MSa is a current, and KW/JB are former (within last 3 years) employees of the Australian Centre for the Prevention of Cervical Cancer (ACPCC) who have received donated laboratory tests, equipment and funding for self-collection related research. LW, LM, RG, JB, TB, MSm, MSa are investigators on a MRFF RART funded project (PACE) which receives donated HPV point-of-care tests from Cepheid. BL reports receiving fees (paid to institution) and holding unpaid positions on the NZ National Cervical Screening Programme Clinical Practice Guidelines Committee and Advisory and Action Group.

## Abbreviations

ACCHO: Aboriginal Community Controlled Health Organisation
AIATSIS: Australian Institute of Aboriginal and Torres Strait Islander Studies
AHMRC: Aboriginal Health and Medical Research Council of NSW
AMSANT: Aboriginal Medical Services Alliance Northern Territory
HPV: Human Papillomavirus
NACCHO: National Aboriginal Community Controlled Health Organisation
NCSP: Australia’s National Cervical Screening Program
NCSR: National Cervical Screening Register
NSW: New South Wales
NT: Northern Territory
QAIHC: Queensland Aboriginal and Islander Health Council
Qld: Queensland

## Notes

### Clinical Trial

Retrospectively Registered with the Australian New Zealand Clinical Trials Registry on 16/10/2025. ACTRN12625001134415 (https://anzctr.org.au/ACTRN12625001134415.aspx)

### Funding Statement

Yes

